# Machine Learning Approach to Integrate and Analyse Multiomics data to Identify Actionable Biomarkers for Head and Neck Squamous Cell Carcinoma (HNSCC)

**DOI:** 10.1101/2025.10.09.25335922

**Authors:** Kajal Panchal, Kulandai Arockia Rajesh Packiam, Sharmistha Majumdar

## Abstract

Head and neck squamous cell carcinoma (HNSCC) is ranked sixth among all the common cancers worldwide and is a major cause of death. A molecular understanding of disease progression can aid in timely diagnosis and therapy. This study aims to identify potential HNSCC biomarkers using a machine learning-based approach to integrate and analyse multi-omics data (namely publicly available Human Papillomavirus (HPV) negative patients’ multiomics datasets from the CPTAC-HNSCC project, including transcriptomics, methylomics, proteomics, and phosphoproteomics). A three-step feature selection method was utilized to identify potential molecular biomarkers using machine learning algorithms. The top 1000 important features (genes) were filtered using Mutual Information, followed by a random forest-based feature importance ranking, and Recursive Feature Elimination with cross-validation coupled with Support Vector Machine (SVM-RFECV) to get a minimal gene set important for machine learning based tumor-normal classification task. To benchmark these top-selected features, Logistic Regression (LogR), Random Forest (RF), Multi-layer perceptron (MLP), and Support Vector Machines (SVC) were used. The prediction performance of classifiers trained on these selected gene sets was evaluated using the accuracy metric, which was then compared against that of models trained on randomly selected gene sets. The entire workflow was repeated 100 times for different random states to establish statistical confidence in the pipeline and the selected gene set. Our integrative approach identified both omics-specific and cross-omics candidate genes with very high classification accuracy, ranging from ∼ 95% to 100%. These genes reveal convergent biological processes central to HNSCC pathogenesis, which reinforces the robustness of the methodology used, which can be adopted to analyse similar multiomics datasets for other pathologies and foundational biological questions.

## 1. Introduction

Head and neck squamous cell carcinoma (HNSCC) represents a highly heterogeneous malignancy with two major etiological subgroups: human papillomavirus (HPV)-positive and HPV-negative tumors. While HPV-positive HNSCC cancers are associated with a better prognosis, higher sensitivity to treatment, and distinct virus-driven biology, HPV-negative HNSCCs are strongly linked to carcinogen exposure such as tobacco and alcohol, and carry significantly worse clinical outcomes with a high recurrence rate and resistance to therapy (Krishnan et al., 2024). Importantly, biomarker development has largely focused on HPV-positive disease, whereas HPV-negative patients continue to face poor prognosis, limited therapeutic options, and lower overall survival (Powell et al., 2021; Waas et al., 2024). Identifying robust biomarkers in HPV-negative HNSCC is therefore a priority to improve precision medicine in this subgroup.

Conventional biomarker discovery strategies have largely focused on single-omics platforms. While such approaches have provided valuable insights into tumor biology, they capture only a fraction of the molecular alterations that drive malignant transformation and progression. Cancers, including HNSCC, arise from complex, multi-layered dysregulation spanning the genome, epigenome, transcriptome, proteome, and post-translational modifications. Therefore, integrative multi-omics strategies are essential to unravel the biological heterogeneity of HNSCC and to identify robust biomarkers that transcend individual molecular layers. Machine learning based algorithms are sensitive to non-linear relationships between biological features and the target class (normal or tumor) across different layers of omics data. Machine learning–based feature selection approaches further empower these analyses, offering scalable solutions to extract relevant features from high-dimensional datasets and to identify signatures with diagnostic, prognostic, or therapeutic relevance.

In the present study, we aimed to capture both omics-specific alterations and cross-platform convergence using the feature importance of machine learning models. Our analysis uncovered recurrent candidates— including secreted serpins, immune regulators, and metabolic enzymes—that not only map to cancer-relevant pathways but also represent promising biomarkers and therapeutic targets. This work underscores the power of integrative multi-omics approaches to refine our understanding of HNSCC biology and to advance precision oncology.

## 2. Materials and Methods

### 2.1 Dataset

HPV-negative HNSCC patient samples were generated by Huang et al. (2021) to understand the proteogenomic depths of HNSCC biological conditions. They collected 109 tumor samples, related blood samples, and 66 matched normal tissue samples under the National Cancer Institute (NCI)’s Clinical Proteomics Tumor Analysis Consortium (CPTAC-3) project. Tumor sites were mainly from the oral cavity and larynx (44.5% each). The authors performed Proteogenomics molecular profiling using high-throughput technologies and uploaded processed and normalized data on LinkedOmics. In this study, the processed data for all the omics was retrieved from the LinkedOmics. Gene-level mRNA data (RSEM) are upper-quantile normalized and log2 transformed. The probe-level methylation data (represented as beta-values) were aggregated to gene-level by extracting probes located in the CpG island of the promoter region of a given gene. The mean beta-value of these probes was used to represent gene-level methylation. Gene-level proteomics data, which were represented as normalized MS intensity, were log2-transformed. Gene-level phosphoproteomics data, which were represented as normalized MS intensity, were log2-transformed

### 2.2 Tools and packages used for the analysis

Open source Python packages, including NumPy(Harris et al., 2020), Pandas (McKinney, 2010), scikit-learn(Pedregosa et al., n.d.), Matplotlib (Hunter, 2007), Seaborn (Waskom, 2021), and SciPy(Virtanen et al., 2020), were used for exploratory data analysis, statistics, visualization, classification tasks, and feature selection steps. The link to the GitHub repository is provided in the supplementary.

### 2.3 Data Pre-processing

Genes with all zeros and NaN values were examined for removal, but none were found. Then, all features with near-zero variance were removed, for which 0.01 was the cut-off variance value for transcriptomics, proteomics, and phosphoproteomics, and 0.001 for methylomics. A total of 2805 and 9389 features got filtered out, respectively, in transcriptomics and methylomics, while none were filtered out in proteomics and phosphoproteomics. In transcriptomics data, 3354 genes with low expression values were removed (features with < 1 log2 RSEM values, which corresponds to < 1 FPKM in 90% of the samples). Transcriptomics data had 0 NaN values, while there were 98 NaN values in Methylomics, Proteomics had 79213 NaN (∼20.98% overall, 21.52% in Normal, 20.9% in Tumor), and phosphoproteomics data had 67810 NaN (∼16% overall, 16.24% Normal, 16.06% in Tumor). Features with > 86 (>50% of the samples) NaN values across samples in proteomics (8 features) and phosphoproteomics (18 features) were removed. After data processing, Transcriptomics was left with 32297 features, 53 (N) + 109 (T) samples, Methylomics with 3728 features and 40 (N) + 104 (T) samples, Proteomics with 9658 features and 63 (N) + 109 (T) samples, Phosphoproteomics with 6342 and 63 (N) + 109 (T) samples, and Multiomics with 52024 features and 28 (N) + 103 (T) samples.

### 2.4 Main Workflow

The overall workflow is shown in Fig.1. The same machine-learning-based workflow, including three step feature selection steps and tumor-normal class prediction tasks, was run for each single omics layer, separately analyzed to select important features(genes) from each molecular layer, after which all four omics layers were concatenated as a single input and subjected to the same workflow for important gene set identification. To avoid any bias due to randomness in the test split, imputation, and model training, the overall workflow was repeated 100 times for 100 different random states. Three feature selection steps reduced the feature set size to get the most important features (here, genes). The outcome of each iteration was the features selected from the SMRFECV step. At the end, the most frequently selected genes across 100 iterations were considered as the final gene set from each single omics layer and from integrated multi-omics analysis. A classification task using four different classifiers gave a measure of the significance of features to differentiate between two classes. At any step, where the random state is a parameter, the random state for the specific iteration was used to maintain reproducibility and uniformity.

**Figure 1.**
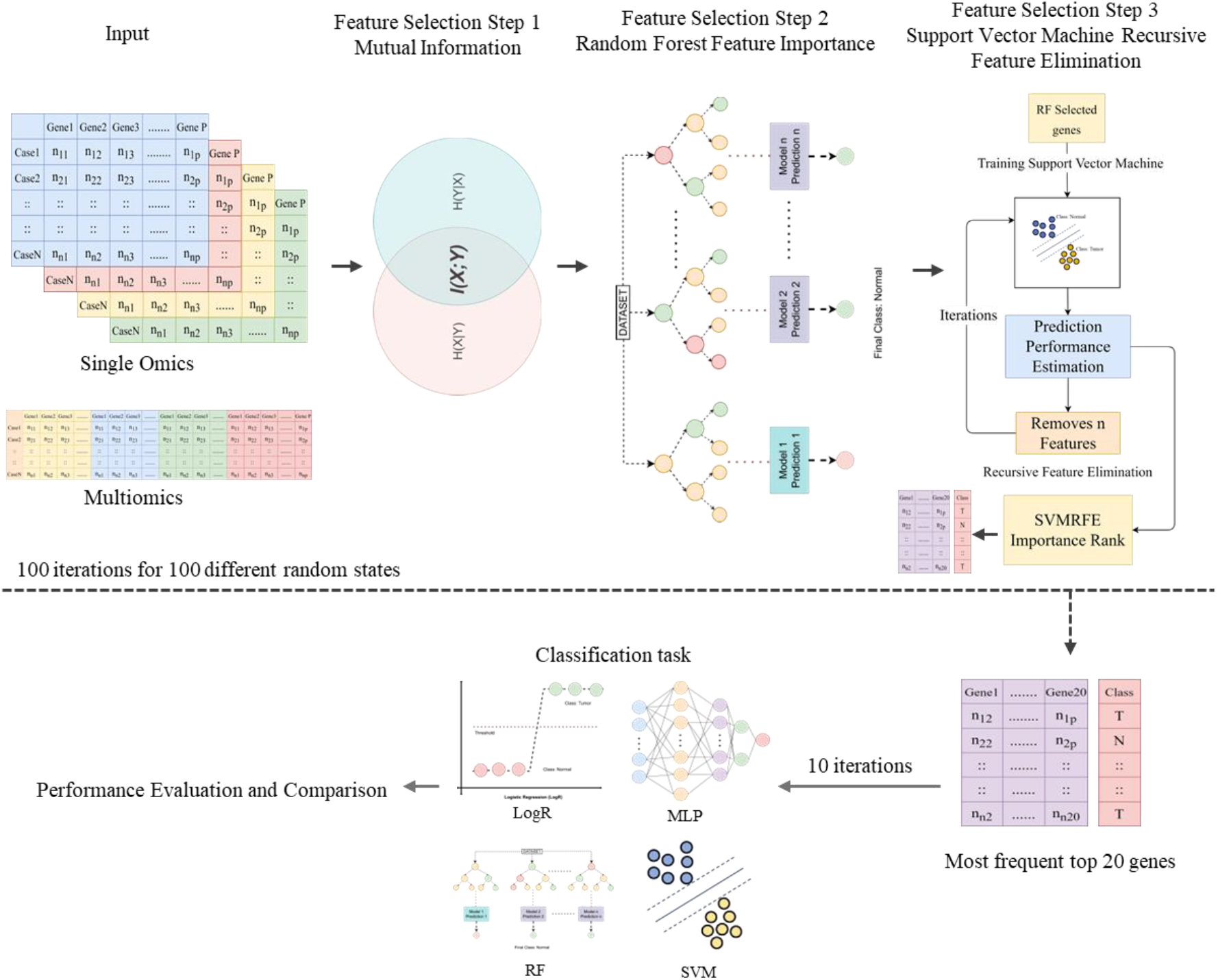
Overall workflow for single omics and multi-omics molecular marker identification strategy.

#### 2.4.1 Train-test split, imputation, and min-max scaling

The data were divided into feature values corresponding to omics layers and target classes. 80% of the data was kept as a training set and 20% as a test set; all the remaining missing values were imputed using the K-nearest neighbours imputer (KNNImputer) from the sklearn impute module. The MinMaxScaler from sklearn was used to get all the values across different features to a similar scale of 0 to 1.

#### 2.4.2 Feature Selection

A three-step gradual feature selection procedure was employed. This approach was considered because of the high-dimensional nature of omics data with thousands of molecular features and a smaller sample size (classic curse of dimensionality). Three steps balance between the computation efficiency and sensitivity to feature association complexity. The core of this selection step is Recurrent Feature Elimination with cross-validation (RFECV), as many research studies have shown its robustness in biomarker identification (Guyon et al., 2002; Sanz et al., 2018; Sun et al., 2023), but it is computationally challenging to fish out a few important ones out of thousands of features at one go using RFECV. Hence, we have added two additional filtering steps before the RFECV step: a mutual information-based step and a Random Forest (RF) based step. We used the mutual_info_classif function from scikit-learn to calculate mutual information, which is a univariate method that captures gene-class relationships beyond linear relationships. Mutual Information filtering reduces the dimensionality by retaining the most informative 1000 features. In the second step, sklearn’s SelectFromModel was used, which ranks features based on the model’s inherent importance score, like in tree-based methods or coefficients in SVM or Logistic Regression. This meta transformer keeps only the most important features by applying a threshold. Here, Random Forest + SelectFromModel was used to identify high-importance features with an importance score more than the threshold (median of all feature importance scores). The max_feature parameter was set to 100 to select genes ranked above 100 or fewer. In the last step, we employed RFECV using sklearn’s RFECV with Support Vector Machine classifier (SVM-RFECV)(Sanz et al., 2018) with five-fold cross-validation. This method systematically prunes less informative features and trains an SVM classifier iteratively to find feature importance on the updated feature set.

#### 2.4.3 Classification task

Logistic Regression (LogR), Random Forest (RF), Support Vector Machine (SVM), and Multi-Layer Perceptron (MLP) were used to perform the classification task using the selected feature set, as well as a random set within every random state iteration (total 100). The same was done after selecting the most frequent significant features for 10 random states to check the classification performance of the final gene set for all four single omics and multiomics.

#### 2.4.4 Statistical Analysis

The robustness of the workflow in picking the most significant genes from the pool of thousands of genes was tested using a statistical test to check if the classification performance of models trained on selected genes was significantly better than that of the model trained on randomly selected genes. We performed the Wilcoxon signed rank test (Rainio et al., 2024) to compare the corresponding model accuracies for both gene sets.

## 3. Results and Discussion

The genes selected by our workflow have been listed in Supplementary Table 1, along with a detailed description of each gene gathered from Human Protein Atlas (Thul & Lindskog, 2018) in a linked sheet. Transcriptomic signatures were dominated by extracellular matrix remodeling genes such as MMP11 and MMP12. Interestingly, neuronal and glial genes, including PLP1, ASPA, and NXPH3, were also dysregulated, pointing to a potential link with perineural invasion, a hallmark of advanced HNSCC. Methylomic features primarily mapped to neuronal development and synaptic signaling (GRM6, GRIN2B, AATK, TAC1), suggesting that epigenetic silencing of neuro-associated pathways may facilitate tumor plasticity. These results underscore the role of DNA methylation in reprogramming developmental circuits during tumor progression. Proteomic data revealed metabolic enzymes (ADH1B, ADH4, CYP4F12), cytoskeletal regulators (KRT13, CAVIN2), and acute-phase/serpin family proteins (SERPINA1, SERPINA6, HPX). These reflect metabolic rewiring and immune modulation, both central to HNSCC biology. Notably, several of these proteins are secreted and detectable in biofluids, making them promising candidates for non-invasive biomarkers. Phosphoproteomic signatures highlighted immune signaling (IFI35, ZBTB7B), cytoskeletal remodeling (CTTNBP2, KRT13), and translational control (EIF4EBP2). These findings suggest that post-translational regulation orchestrates both tumor–immune interactions and dynamic cytoskeletal changes.

**Figure 2.**
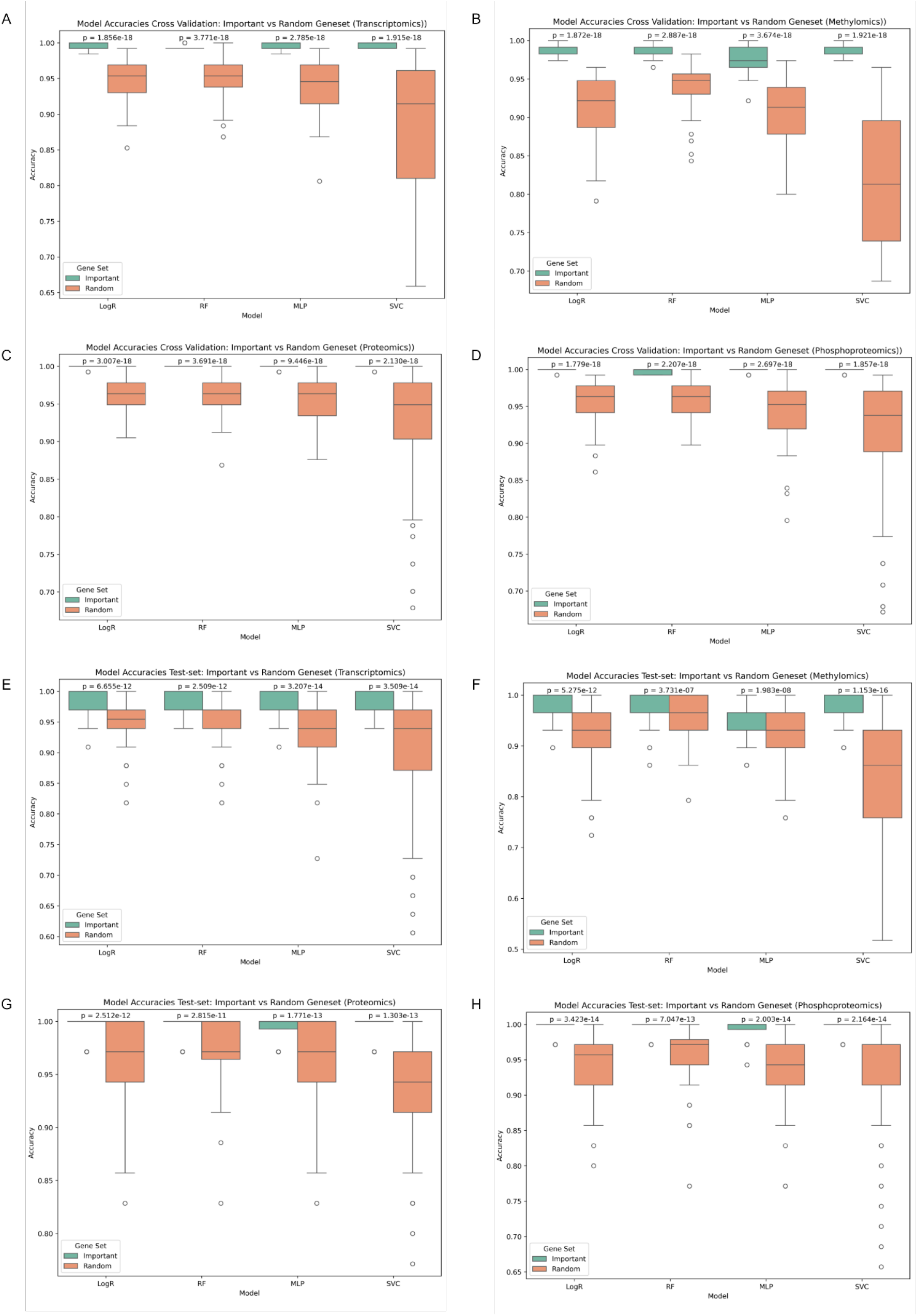
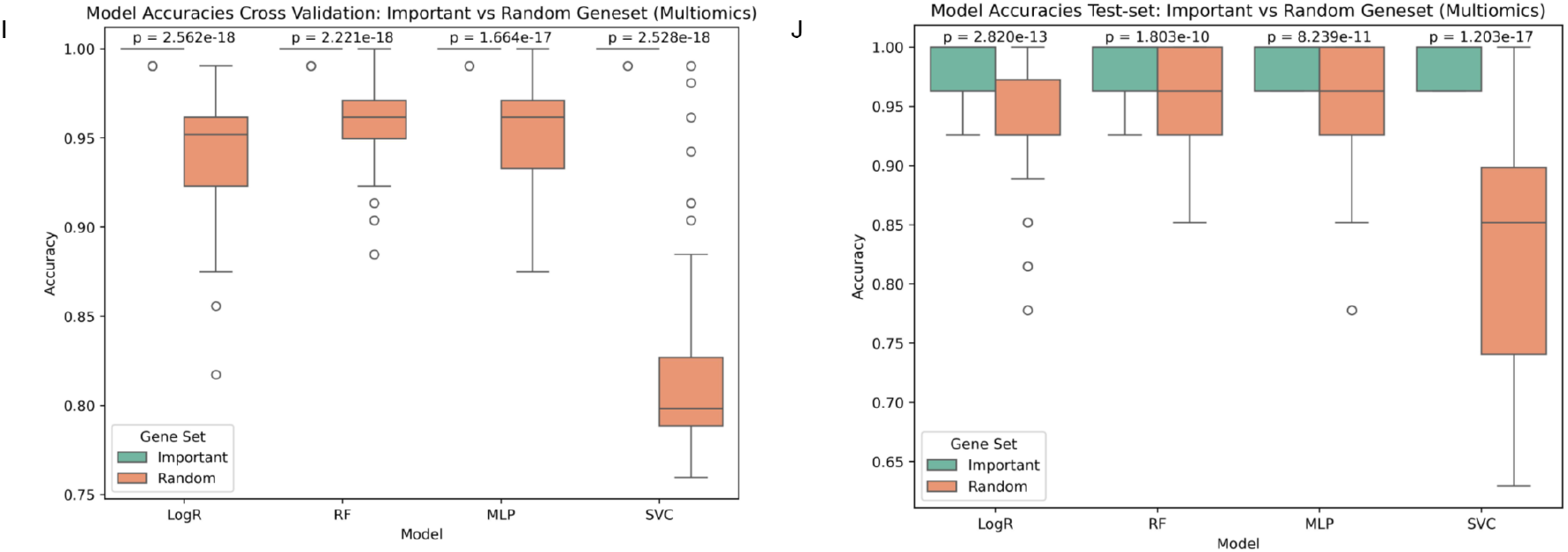
Cross-validation and prediction accuracy distribution for different classifiers across different omics analyses (100 random state iterations). p-values are calculated for the Wilcoxon signed rank test (alternative hypothesis: the accuracy of classifiers trained on selected geneset is greater than the models trained on a random geneset).

When all omics layers were combined, the consensus multiomics signature strongly converged on secreted protease inhibitors (SERPINA1, SERPINA3, SERPINA6, SERPINA7, SERPING1), acute-phase proteins (HPX, ORM2, AGT), and immune regulators (IFI35). This indicates that HNSCC tumors engage in profound remodeling of their extracellular and immune microenvironment, with systemic consequences detectable in circulation. Several genes were consistently highlighted across multiple layers, including SH3BGRL2 (transcriptome, proteome, multiomics), GREM2 (transcriptome, phosphoproteome), CAVIN2 (proteome, phosphoproteome), IFI35 (phosphoproteome, multiomics), TRPM6 (phosphoproteome, multiomics), and KRT13 (proteome, phosphoproteome).

These multi-omics concordant genes are strong candidates for functional validation and biomarker development. Their presence across RNA, protein, and post-translational levels suggests that they represent stable biological alterations rather than noise from a single platform. The machine learning pipeline has successfully identified a rich list of gene candidates that are central to HNSCC pathobiology. The findings underscore the value of moving beyond single-omics studies to achieve a more holistic understanding of this complex disease. The enrichment of secreted acute-phase proteins and serpins in the consensus signature further supports their potential as blood-based biomarkers for diagnosis or prognosis. Accuracy values in Table 1 for the final gene set and the p-values from the Wilcoxon signed-rank test for accuracy distribution over 100 random states show that the models trained on the selected gene sets perform significantly better than the models trained on a random gene set, which supports the robustness of the strategy to find potential molecular markers. Overall, our approach offers a transferable strategy for leveraging multi-omics data to advance mechanistic insights and therapeutic discovery in complex diseases.

**Table 1.** Mean accuracy and standard deviation of classifiers trained on the final geneset from the most frequently occurring significant genes after three-step feature selection steps over 100 iterations of random states.

| Models | Transcriptomics | Methylomics | Proteomics | Phosphoproteomics | Multiomics |
| --- | --- | --- | --- | --- | --- |
| LogR | 0.997 ± 0.01 | 0.993 ± 0.015 | 0.991 ± 0.014 | 1.0 ± 0.0 | 1.0 ± 0.0 |
| RF | 0.997 ± 0.01 | 0.997 ± 0.011 | 1.0 ± 0.0 | 0.991 ± 0.014 | 1.0 ± 0.0 |
| MLP | 0.997 ± 0.01 | 0.952 ± 0.044 | 0.991 ± 0.014 | 1.0 ± 0.0 | 1.0 ± 0.0 |
| SVC | 0.997 ± 0.01 | 0.983 ± 0.018 | 1.0 ± 0.0 | 1.0 ± 0.0 | 1.0 ± 0.0 |

## Supporting information

Supplemental section

## Data Availability

https://github.com/kajalpanchal314/HNSCC_Multiomics

## Notes

### Competing Interest Statement

The authors have declared no competing interest.

### Funding Statement

GSBTM

### Author Declarations

The study used (or will use) ONLY openly available human data that were originally located at CPTAC

