## Supplemental section for "Machine Learning Approach to Integrate and Analyse Multiomics data to Identify Actionable Biomarkers for Head and Neck Squamous Cell Carcinoma (HNSCC)"

#### Supplementary Information

| Sr. No. | Transcriptomics | Methylomics | Proteomics | Phospho-proteomics | Multiomics |
|---|---|---|---|---|---|
| 1 | GREM2 | GRM6 | ADH4 | IFI35 | P_HPX |
| 2 | SASH1 | FOXR1 | SH3BGRL2 | SKI | P_SERPINA6 |
| 3 | CAB39L | GPR75 | SCIN | CAVIN2 | T_ETFDH |
| 4 | CLEC3B | VSX1 | PID1 | GREM2 | P_NOSTRIN |
| 5 | ASPA | NLRP4 | CYP4F12 | CTTNBP2 | P_SERPINA1 |
| 6 | F10 | SMPD3 | ADAR | EVA1C | P_SH3BGRL2 |
| 7 | LOC105373265 | CCDC181 | HPX | TPPP | P_ORM2 |
| 8 | ETFDH | AATK | CAVIN2 | PLEKHA7 | P_SERPING1 |
| 9 | MMP11 | VSTM2A | KRT13 | OXR1 | P_SERPINA7 |
| 10 | MMP12 | COX8C | C2orf54 | ZBTB7B | T_ASPA |
| 11 | TMEM132C | SPIN2A | SERPINA6 | SERPIND1 | PP_IFI35 |
| 12 | NRG2 | NT5DC3 | ADH1B | TRPM6 | P_SERPINA3 |
| 13 | PLP1 | GRIN2B | SERPINA1 | AHSG | T_LOC105373265 |
| 14 | SH3BGRL2 | ABCC9 | STAT2 | KNG1 | P_PID1 |
| 15 | GGTA1P | FOXD2 | IL33 | SRSF2 | T_ADAM12 |
| 16 | CBX3 | RGN | FNDC3B | KRT13 | P_LYVE1 |
| 17 | KIF26B | UBBP4 | GTPBP4 | CFI | T_QARS |
| 18 | NXPH3 | TAC1 | UACA | PLCD1 | T_MMRN1 |
| 19 | MINDY1 | ADAMTS18 | ATP6V1C1 | EIF4EBP2 | PP_TRPM6 |
| 20 | CTTNBP2 | KBTBD12 | BCKDHA | FAM129B | P_AGT |

Supplementary Table 1. Final gene list of the most significant genes (prefix in multiomics represents the origin of the omics layer of the feature)

More information on each selected gene (collected from Human Protein Atlas): [Link to Excel sheet]

Overlapping important features

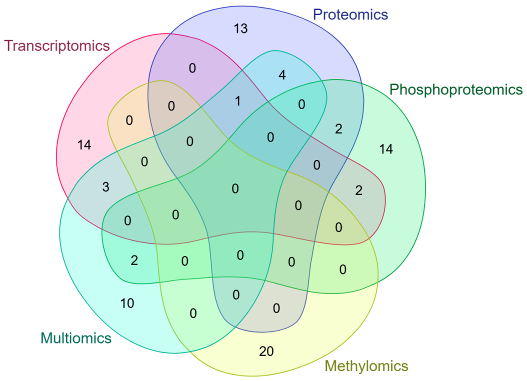


#### Shared Items

| Item | Occ. | Present in |
|---|---|---|
| 'SH3BGRL2' | 3 | Transcriptomics, Proteomics, Multiomics |
| 'ASPA' | 2 | Transcriptomics, Multiomics |
| 'CAVIN2' | 2 | Proteomics, Phosphoproteomics |
| 'CTTNBP2' | 2 | Transcriptomics, Phosphoproteomics |
| 'ETFDH' | 2 | Transcriptomics, Multiomics |
| 'GREM2' | 2 | Transcriptomics, Phosphoproteomics |
| 'HPX' | 2 | Proteomics, Multiomics |
| 'IFI35' | 2 | Phosphoproteomics, Multiomics |
| 'KRT13' | 2 | Proteomics, Phosphoproteomics |
| 'LOC105373265' | 2 | Transcriptomics, Multiomics |
| 'PID1' | 2 | Proteomics, Multiomics |
| 'SERPINA1' | 2 | Proteomics, Multiomics |
| 'SERPINA6' | 2 | Proteomics, Multiomics |
| 'TRPM6' | 2 | Phosphoproteomics, Multiomics |

### Variance information of features

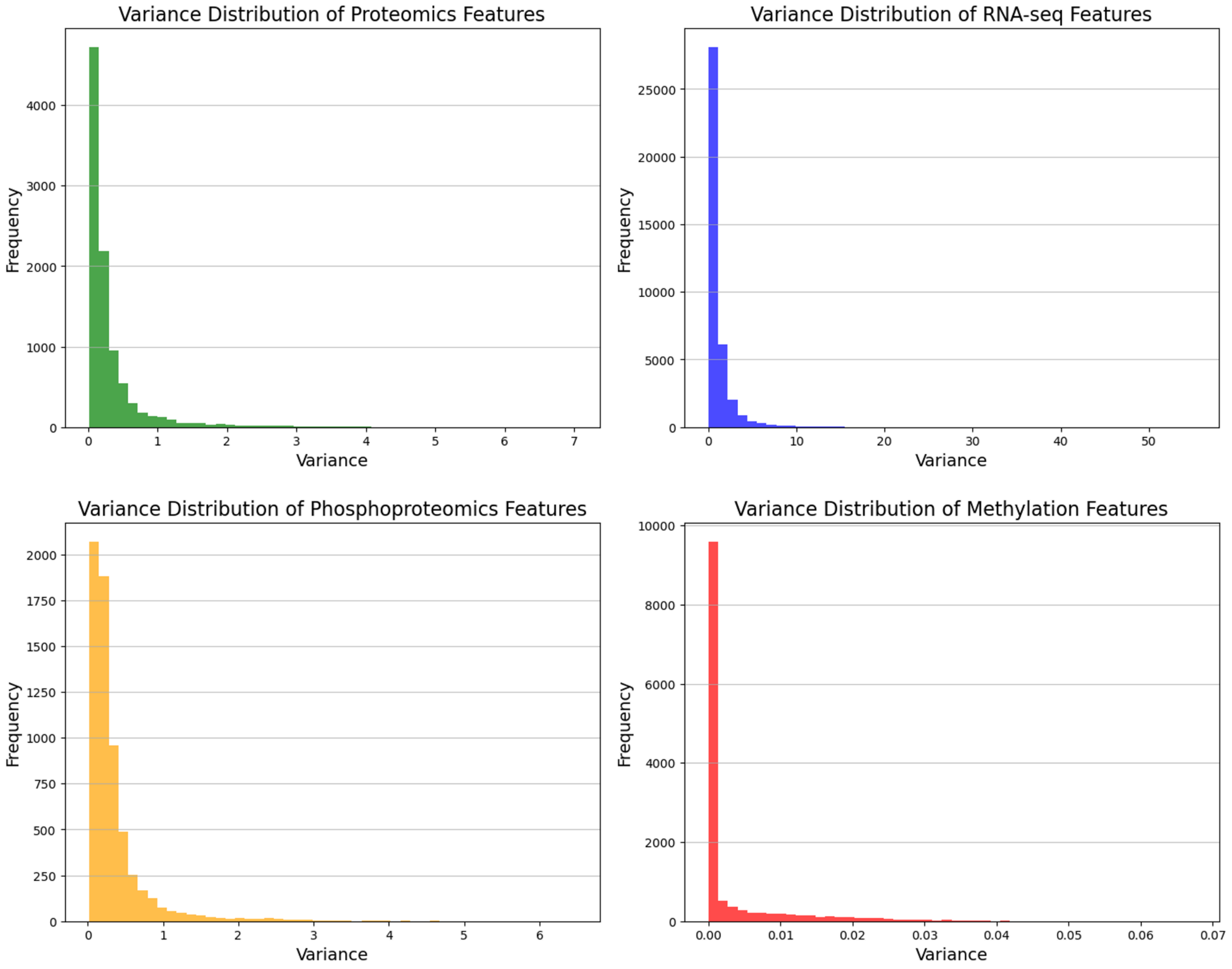

Number of features selected in the Random Forest-based and SVMRFECV-based selection steps across 100 iterations of random states

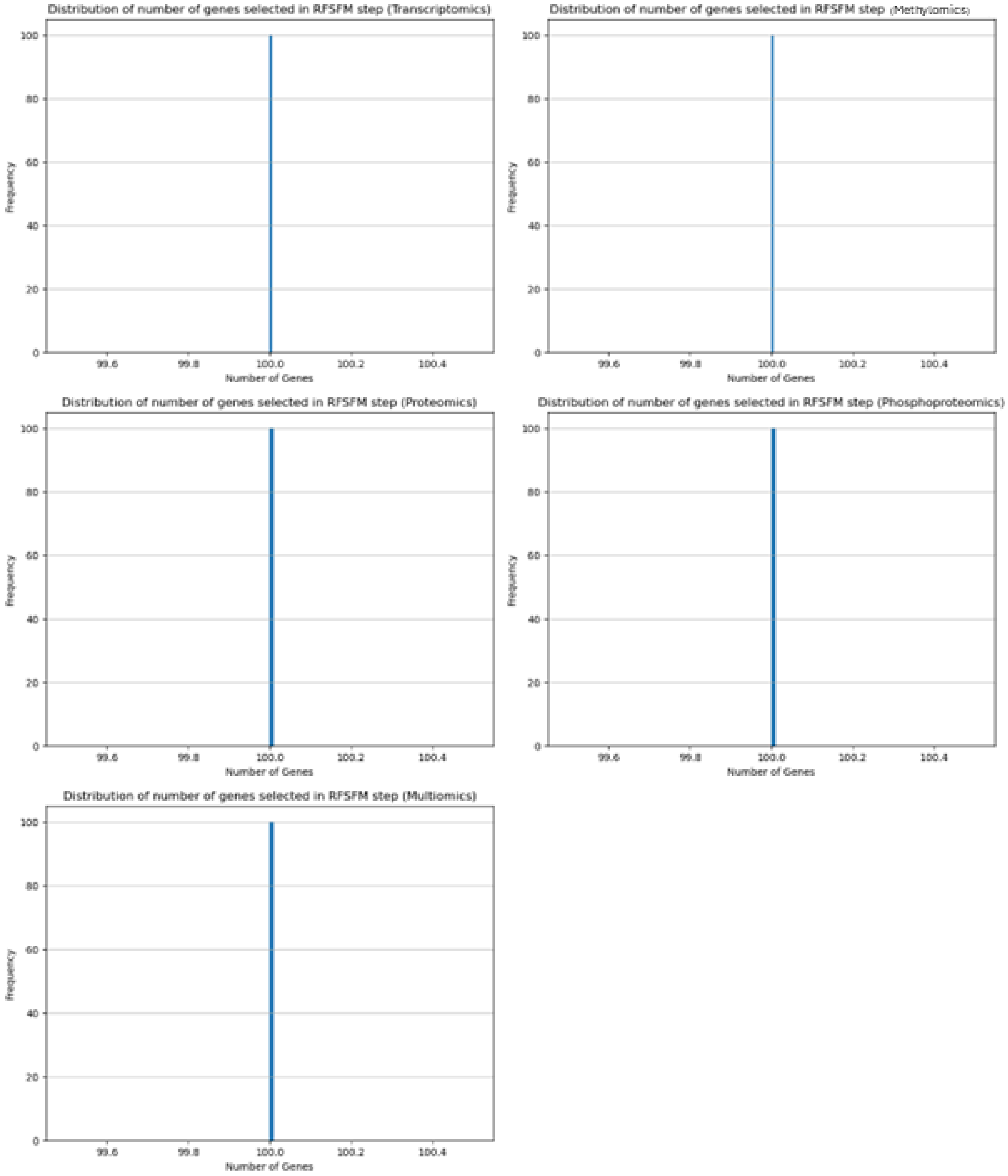

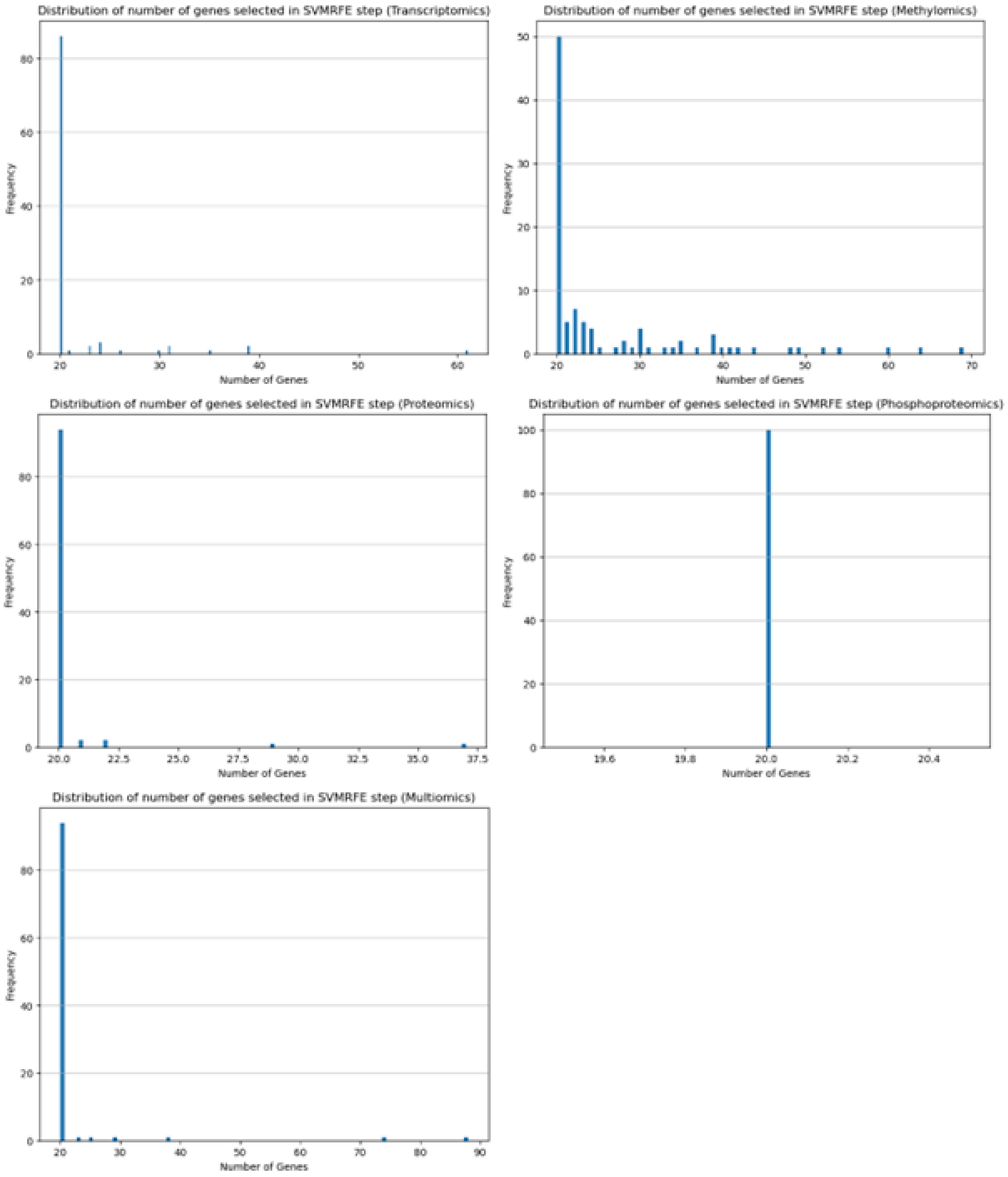


Code availability: https://github.com/kajalpanchal314/HNSCC_Multiomics
